# Identification of probable child maltreatment using prospectively recorded information between 5 months and 17 years in a longitudinal cohort of Canadian children

**DOI:** 10.1101/2023.04.05.23288127

**Authors:** Sara Scardera, Rachel Langevin, Delphine Collin-Vezina, Maude Comtois Cabana, Snehal M Pinto Pereira, Sylvana Côté, Isabelle Ouellet-Morin, Marie-Claude Geoffroy

**Affiliations:** Department of Educational and Counselling Psychology, McGill University, Education Building, 3700 McTavish Street, H3A 1Y2, Montreal, Quebec, Canada; School of Social Work, McGill University, Montreal, Quebec, Canada; Department of Psychology, University of Montreal, Montreal, Quebec, Canada; Faculty of Health Sciences, Simon Fraser University, British Columbia, Canada; Faculty of Medical Sciences, University College London, England, United Kingdom; Department of Social and Preventive Medicine, University of Montreal, Montreal, Quebec, Canada; School of Criminology, University of Montreal & the Research Center of the Montreal Mental Health University Institute, Montreal, Quebec, Canada; McGill Group for Suicide Studies, Douglas Mental Health University Institute & Department of Psychiatry, McGill University, Montreal, Canada

**Author notes:** **Corresponding Author:** Marie-Claude Geoffroy, PhD, Department of Educational and Counselling Psychology, McGill University, Montreal, QC H3A 1Y2, Canada. Drs. Ouellet-Morin and Geoffroy shared senior authorship. **Role of the Funder/Sponsor:** The funder had no role in the design and conduct of the study; collection, management, analysis, and interpretation of the data; preparation, review, or approval of the manuscript; and decision to submit the manuscript for publication.

**Keywords:** Child maltreatment, measurement, population-based cohorts, methodology

## Abstract

**Background:** Both prospective and retrospective measures of child maltreatment predict mental health problems, despite their weak concordance. Research remains largely based on retrospective reports spanning the entire childhood due to a scarcity of prospectively completed questionnaires targeting maltreatment specifically.

**Objective:** We developed a prospective index of child maltreatment in the Québec Longitudinal Study of Child Development (QLSCD) using prospective information collected from ages 5 months to 17 years and examined its concordance with retrospective maltreatment.

**Participants and Setting:** The QLSCD is an ongoing population-based cohort that includes 2,120 participants born from 1997-1998 in the Canadian Province of Quebec.

**Methods:** As the QLSCD did not have maltreatment as a focal variable, we screened 29,600 items completed by multiple informants (mothers, children, teachers, home observations) across 14 measurement points (0-17 years). Items that could reflect maltreatment were first extracted. Two maltreatment experts reviewed these items for inclusion and determined cut-offs for possible child maltreatment. Retrospective maltreatment was self-reported at 23 years.

**Results:** Indicators were derived across preschool, school-age and adolescence periods and by the end of childhood and adolescence, including presence (yes/no), chronicity (re-occurrence), extent of exposure and cumulative maltreatment. Across all developmental periods, the presence of maltreatment was as follows: physical abuse (16.3-21.8%), psychological abuse (3.3-21.9%), emotional neglect (20.4-21.6%), physical neglect (15.0-22.3%), supervisory neglect (25.8-44.9%), family violence (4.1-11.2%) and sexual abuse (9.5% in adolescence only).

**Conclusions:** In addition to the many future research opportunities offered by these prospective indicators of maltreatment, this study offers a roadmap to researchers wishing to undertake a similar task.

**Highlights:**

- In this longitudinal cohort, maltreatment experts retained 251 of 29,600 items available
- Probable maltreatment indicators were derived: presence, chronicity, extent of exposure, and cumulative maltreatment
- Prevalence rates vary from 3.3% and 44.9% across developmental periods, and 16.5-67.3% by the end of adolescence
- Prospective and retrospective maltreatment identify different groups of individuals
- As most studies use retrospective data, findings suggest that the representation of child maltreatment is incomplete and retrospective reports should be complimented by prospective data, whenever possible

## Introduction

Child maltreatment refers to “any act or series of acts of commission or omission by a parent or other caregiver that results in harm, potential for harm, or threat of harm to a child” (Arias et al., 2008, p. 11). Maltreatment increases the risk for a range of physical health (e.g., obesity, cardiovascular diseases) and mental health problems (e.g., suicide attempts, depression and substance use) across the lifespan (Gilbert et al., 2009; Jaffee, 2017; Min et al., 2013; Nanni et al., 2012). Despite strong evidence supporting the harmful consequences of abuse and neglect on later functioning, the field continues to face its biggest methodological challenge: the very measurement of child maltreatment (Danese & Widom, 2020; Shaffer et al., 2008). Obtaining accurate assessment of maltreatment is not straightforward given limitations noted across all measurement strategies, and resulting wide ranges in estimates depending on the source of information (e.g., official vs. retrospective reports) (Gilbert et al., 2009). For example, official records (maltreatment records documented by youth protection services) are hampered by under reporting and may only capture the most severe cases (Jaffee, 2017), whereas questionnaires completed by caregivers may be subjected to social desirability (Fisher & Katz, 2000). While retrospective self-reports, completed by the targeted participants, is less prone to social desirability than those filled out by caregivers, they may be more affected by current mental health (e.g., depressive symptoms) or memory accuracy (Danese & McCrory, 2015). Research has shown that prospective and retrospective reports of maltreatment are associated with mental health outcomes (albeit to different extents), and identify different groups of individuals (Baldwin et al., 2019; Danese & Widom, 2020). As such, it may be advised to also consider prospectively collected indicators of maltreatment, collected across multiple informants, to widen our representation of childhood experiences of maltreatment beyond retrospective measures and official records. This study aims to describe the creation of prospective indicators of maltreatment, building on all available information collected in the population-based Quebec Longitudinal Study of Child Development (QLSCD) cohort, from the time participants were 5 months old up to 17 years of age.

### Measuring probable maltreatment using prospectively collected information

To our knowledge, only few population-based longitudinal cohorts have prospectively measured child maltreatment (Denholm et al., 2013; Houtepen et al., 2018; Kisely et al., 2020; Naicker et al., 2022; Newbury et al., 2018; Patten et al., 2015; Reuben et al., 2016). Prospective information can be obtained, for instance, from Youth Protection official records of notified or substantiated maltreatment or through direct questions to caregivers or participants themselves using standardized questionnaires or interviews (e.g., structured interview about child harm during home visits (Newbury et al., 2018)). Although official records and prospectively collected caregiver information are valuable, especially when complimented by retrospective self-reports, they remain rare, especially spanning several periods of development.

Although there is no gold standard approach for collecting maltreatment data, prospective longitudinal cohorts offer additional opportunity to derive indicators of probable maltreatment (proxy) using general items (non-specific to maltreatment) and data collected across multiple informants and developmental stages. For instance, in the Avon Longitudinal Study of Child Development (ALSPAC) (Houtepen et al., 2018), an adversity index encompassing the ten classic adverse childhood experiences (ACEs) (Felitti et al., 1998) was derived using 541 prospective items responded by parents and children (> 8 years) collected from birth to 18 years. Although most response options were in frequencies (e.g., never to everyday), cut-offs were used to dichotomize each item. Two variables were created, including the presence of distinct types of adversity and a cumulative score (i.e., sum of the types of adversity an individual was exposed to). These derived adversity variables have since been associated with increased depression and drug use in adolescents (Houtepen et al., 2020). Using a similar procedure, an indicator of neglect, operationalized by two variables (presence and severity), was derived in the 1958 British Birth cohort using seven items administered to mothers, fathers, and teachers at seven, 11, and 16 years (Denholm et al., 2013). This indicator has been associated with mental health, cognition, and obesity in adult life (Degli Esposti et al., 2020; Geoffroy et al., 2016; Power et al., 2015), even after controlling for key socioeconomic confounding factors. Typically, these longitudinal cohorts offer global indicators of maltreatment (e.g., presence versus absence) and consider the lifetime occurrence of maltreatment (e.g., any time from birth to 18 years). However, more specific characteristics of maltreatment or adversity (e.g., chronicity), as well as the specific time of occurrence of these experiences are often overlooked.

### Importance of research on specific characteristics of child maltreatment

Research suggests that child maltreatment is multidimensional in nature, and several characteristics of maltreatment may jointly contribute to explain later risk for specific mental health difficulties (Cicchetti & Toth, 2005; Egeland et al., 1983; Jackson et al., 2019). Yet, limitations remain as child maltreatment has typically been operationalized through global conceptualizations (presence versus absence of child maltreatment) or by a single type of abuse (e.g., physical or sexual) or neglect. Although challenging, important characteristics of maltreatment should be simultaneously considered to investigate their common and specific contributions, as outlined below.

### Type

The most common dimension for operationalizing child maltreatment is through the categorization of distinct types (e.g., physical abuse, sexual abuse, emotional neglect) (Jackson et al., 2019). Studies suggest that individual types of maltreatment may contribute specifically or in a shared manner to later outcomes (Cecil et al., 2017; Cheng & Langevin, 2022). For example, a study of emerging adults found that a history of emotional abuse contributed globally to the dimensions of emotional regulation, whereas other types of maltreatment (e.g., neglect) contributed individually to specific facets of emotional regulation (e.g., impulsivity) (Cheng & Langevin, 2022). Additionally, most research on child maltreatment and later outcomes has focused on physical and sexual abuse (Angelakis et al., 2019; Baldwin et al., 2019; Norman et al., 2012). Conversely, other types of maltreatment have been understudied, including neglect (Stoltenborgh et al., 2013) and psychological abuse (Jackson et al., 2019). As such, studies that provide information on the wider breath of maltreatment types can allow for more insight on the relative effects of each maltreatment type, as well as their combination.

### Cumulative scores

Maltreatment types are highly correlated and often co-occur (Kessler et al., 2010). Despite evidence for individual types being differentially associated with outcomes, growing evidence shows that the number of maltreatment types an individual was exposed to, relates to poorer outcomes later in life (Gilbert et al., 2009; Putnam et al., 2013). For instance, evidence shows a dose-response relation between cumulative maltreatment exposure and more severe symptomology, including heightened risk for suicide ideation and self-harm (Turner & Colburn, 2022) as well as anxiety and depression (Finkelhor et al., 2007). Yet, most studies do not consider the cumulative effects of child maltreatment and tend to focus exclusively on one maltreatment type (e.g., physical abuse). Consequently, associations between specific maltreatment types and outcomes may be overestimated on their own and underestimated in conjunction with co-occurring types of maltreatment.

### Recurrence, chronicity, and developmental timing

Child maltreatment can be transient (e.g., situational or limited in time) or it can reoccur over time and over several developmental periods. Developmental chronicity of maltreatment (Manly, 2005) is an important characteristic to consider to adequately ascertain the consequences of maltreatment on functioning across the lifespan. Studies have found that exposure to maltreatment over several developmental stages poses a higher risk for the onset of mental health problems compared to exposure at one developmental period (Jaffee & Maikovich_JFong, 2011; Russotti et al., 2021; Thornberry et al., 2001; Warmingham et al., 2019).

Moreover, the timing of exposure (e.g., whether maltreatment occurred in preschool versus school-age versus adolescence) can also provide specificity regarding differential outcomes. Based on substantiated reports of sexual abuse, physical abuse and neglect, Thornberry et al. (2010) found that individuals exposed to any type of maltreatment during childhood were more likely to report internalizing problems (i.e., suicidal thoughts and depression) in early adulthood, while those who were exposed later on, in adolescence, were more likely to exhibit externalizing problems (e.g. criminal behavior and substance use (Thornberry et al., 2010). Another study found that maltreatment occurring earlier in life (e.g., infancy and toddlerhood) was more strongly associated with poor emotion regulation in childhood (Kim & Cicchetti, 2010), than maltreatment occurring later in preschool/school-age. Developmental chronicity and timing can be more challenging to capture in comparison to global indicators (i.e., presence versus absence) to ascertain that maltreatment of a similar type persists rather than swapped by experienced of another type, contributing to a loss of acuity in subsequent analyses. To our knowledge, there are no population-based longitudinal cohorts that consider chronicity and timing, in addition to other maltreatment-based characteristics despite their longitudinal design.

### The present study

The study of child maltreatment is complex, as all experiences are unique to some degree. Longitudinal study designs can allow for the consideration of time-variant maltreatment indicators and patterns. Using prospectively collected data from a large population-based cohort, the Quebec Longitudinal Study of Child Development (QLSCD), we hereby describe the process implemented to derive multiple prospective indicators of child maltreatment during three developmental periods (preschool, school-age and adolescence) and by the end of childhood (birth to 12 years) and adolescence (birth to 17 years). Specifically, we first provided a roadmap for the derivation of the following variables: (a) the *probable presence* of seven types of maltreatment (i.e., sexual, physical and psychological abuse, family violence, and emotional, physical, and supervisory/educational neglect), and (b) the scores of *cumulative maltreatment* referring to the number of types of maltreatment experienced at each developmental period and by the end of childhood and adolescence. Second, we described how other indicators relevant in child maltreatment research could be derived to complement the above-described indices, including (c) *maltreatment recurrence* and *chronicity* (repeated occurrence of each type of maltreatment within and across developmental periods, respectively). In an exploratory fashion, we derived the (d) *extent of exposure*, referring to the number of different or repeating acts. Third, we compared the prevalence resulting from prospective and retrospective assessments of maltreatment and examined the level of concordance between these measures.

## Method

### Definition of child maltreatment

The following seven maltreatment categories of child maltreatment were selected for inclusion: sexual abuse, (2) physical abuse, (3) psychological abuse, (4) emotional neglect, (5) physical neglect, (6) exposure or presence of family violence, and (7) supervisory/educational neglect. These categories and their definitions, presented in **Table 1**, are in accordance with the Québec Youth Protection Act (Québec, 2021) and the Québec Directors of Youth Protection (*Grounds for Reporting a Situation*, 2022). These are also aligned with international definitions of child maltreatment (e.g., the Center for Disease Control and Prevention) (Arias et al., 2008).

**Table 1.** Definitions of probable child maltreatment.

| **Maltreatment types** | **Definitions** |
| --- | --- |
| Physical abuse | A situation in which the child is the victim of bodily injury or is subjected to unreasonable methods of upbringing by his parents or another person, and the child's parents fail to take the necessary steps to put an end to the situation. |
| Sexual abuse | The child has been subjected to acts sexual in nature by the child's parents or another person, with or without physical contact. |
| Psychological abuse | A child is seriously or repeatedly subjected to behaviour on the part of the child's parents or another person that could cause harm to the child, and the child's parents fail to take the necessary steps to put an end to the situation (e.g., denigration, emotional rejection, excessive control, threats). |
| Family (indirect) violence | Children are, in these cases, exposed to domestic or family violence. A child may witness violent words or gestures between their parents, or at the place of another family member. The child may also be exposed to severe separation conflicts. |
| Emotional neglect | Acts of omission, another form of direct ill-treatment, usually manifest themselves in a parent's lingering indifference to their child. A coldness and lack of investment in the parent-child relationship is palpable. The parent is considerably lacking in emotional sensitivity towards their child. |
| Physical neglect | Failing to meet the child's basic physical needs with respect to food, clothing, hygiene, or lodging, taking into account their resources. |
| Educational neglect/supervisory neglect | Failing to provide the child with the appropriate supervision or support or failing to take the necessary steps to ensure that the child receives proper education and stimulation, and if applicable, that he attends school as required under the Quebec Education Act or any other applicable legislation. |

*Note.* Extracted from the Quebec Youth Protection online sources; extended definitions and examples can be found online (Youth Protection Act, 2021; Grounds for Reporting a Situation, 2022).

### Participants and Procedures

The QLSCD (Orri et al., 2021) is an ongoing longitudinal cohort of children born in 1997-1998 between 24 and 42 weeks of gestation to mothers residing in the Canadian province of Québec and speaking either French or English. Families from all regions of Québec were included, excluding administrative regions 10 (Northern Québec), 17 (Cree Territory), 18 (Inuit Territory) (2.2% of all births). The Québec Master Birth Registry of the Ministry of Health and Social Services was used to randomly select participants based on living area and birth rates (Jetté M, 2000; Orri et al., 2021). The final longitudinal cohort included 2120 participants from primarily White European descendants, which was representative of the ethnic distribution in Québec at the cohort’s inception, and initially covered the full range of socioeconomic statuses. To derive the child maltreatment indicators, we used information collected across three developmental periods (1) preschool – six timepoints at 5, 17, 29, 41, 45-56 months and 5 years, (2) school-age – five timepoints at 6, 7, 8, 10, 12 years, and (3) adolescence – three timepoints at 13, 15 and 17 years. Participants also retrospectively reported on their child maltreatment history at age 23 years (see **Supplemental Table 1** for items).

The QLSCD’s data collection is conducted by the Institut de la Statistique du Québec. All the data collected and presented in this study has been approved by ethical committees of Institut de la Statistique du Québec and the CHU Sainte-Justine Hospital Research Centre. The 2021 Special Round data collection (23 years) was also approved by the Douglas Research Center Ethics Committee and by the CHU Ste-Justine research ethics committee. Written informed consent was obtained from participants and-or their parents at each data collection.

### Search strategy

The items search strategy is presented in **Figure 1**. At step 1, all available items between 5 months and 17 years (∼29,600 items) were screened by two independent screeners (SS, MCC) to determine (1) the eligibility of the items, and (2) the appropriate maltreatment categorization (e.g., physical abuse). Information from all informants were considered except fathers’ reports as the rate of missingness was high and uncertainties remained about the frequency of contact between them and their child in instances of parental separation. Thus, their capacity to adequately evaluate the specific experiences enquired in the considered items was questionable (Orri et al., 2021). Four different informants were retained: mothers, teachers, interviewer observations (Bradley & Caldwell, 1977), and the target child. SS compared the lists of items retained by SS and MCC; duplicate items were removed. The following information was extracted for each retained item: child’s age, informant (mother, interviewer observations, child, teacher) and the corresponding maltreatment type.

Maltreatment experts (RL and DCV) then independently reviewed the retained items to evaluate their suitability and determined at which response option each item would be indicative of the presence of maltreatment while considering the developmental period of the child (e.g., never, about once a week or less, a few times a week, one or two times each day, many times each day). Specifically, item selection and determination of cut-off scores were pursued on the basis that a stand-alone item could reflect serious concerns over possible maltreatment. For example, the item “how often do you tell him/her that he/she is bad or not as good as others?” was recoded as “absence” if parents answered “never” or “about once a week or less” and “probable maltreatment” if parents answered “a few times a week” or more at 5 months. However, at 17 months, the item was recoded as “absence” if parents answered “never”, “about once a week or less” or “a few times a week” and “probable maltreatment” when “one or two times each day” or more was endorsed. We opted for a more rigorous cut-off approach, given that certain scales (i.e., 0 [not at all what I did] to 10 [exactly what I did]) lacked definitive clarity regarding the intended measure (e.g., measuring severity versus frequency of the targeted behavior). As such, depending on the positive or negative valence of items, either extreme (0 or 10) of the scale were used as the indication of maltreatment. RL, DCV and SS met to discuss discrepancies and to make final decisions about inclusion and cut-offs. Based on the determined cut-off, all items in the final sample were scored 0 (absence) or 1 (probable maltreatment).

## Statistical Analyses

### Deriving child maltreatment indicators

The individual items retained by the maltreatment experts were used to derive four indicators of child maltreatment: 1. presence by type of maltreatment, 2. cumulative maltreatment, 3. recurrence and chronicity of maltreatment (by type) and, 4. extent of exposure to different or repeating acts. These indicators were derived at each developmental period (preschool, school-age, and adolescence) as well as by the end of childhood (birth to 12 years) and by the end of adolescence (birth to 17 years). The definitions for each indicator along with the coding decisions used to derive each variable are presented in **Table 2**.

**Table 2.** Deriving probable child maltreatment indicators.

|  | According to developmental period | Variables derived by the end of (1) childhood (birth to 12 years) or (2) adolescence (birth to 17 years) |
| --- | --- | --- |
| **Presence by type of maltreatment** | Types of maltreatment experienced (i.e., physical abuse, psychological abuse) scored as “probable maltreatment” (i.e., presence of a given type at any age point<sup>a</sup> within a development period) or “absence” (i.e., calculated only when at least 2/3 of age points were available). | Types of maltreatment experienced (e.g., physical abuse) by the end of childhood (i.e., scored as “probable maltreatment” if a given type was present at preschool and/or school-age) and by adolescence (i.e., scored as “probable maltreatment” if a given type of maltreatment was present at preschool and/or school-age and/or in adolescence). This was derived when all developmental periods for a given type were available (i.e., 2/2 by childhood and 3/3 by adolescence). |
| **Cumulative maltreatment** | Total number of maltreatment types experienced (scored as: 0, 1, 2, 3+). It was derived only when at least 2/3 of the indicators *presence by type of maltreatment* were available within a given developmental period. | Total number of maltreatment types (0, 1, 2, 3+) experienced by the end of childhood (over preschool and school-age) and by adolescence (over preschool, school-age and adolescence). This was calculated only when at least 2/3 of indicators for the *presence by type of maltreatment* were available by the end of childhood or by the end of adolescence.<sup>b</sup> |
| **Recurrence and chronicity, by types of maltreatment** | Total number of times a child was exposed to a type of maltreatment within a developmental period: 'no recurrence (0 or 1 age point)' 1 'recurrence (2+ ages points)'. It was derived when at least 2/3 of the indicators *presence by types of maltreatment* were available within a given developmental period. | Total number of developmental periods (chronicity) a child was exposed to a given maltreatment type. This was derived when all developmental periods were available for a given type (i.e., 2/2 by childhood and 3/3 by adolescence).<br>By childhood: 0, 1 (no recurrence over developmental periods) versus 2+ developmental periods (recurrence).<br>By adolescence: 0, 1 (no recurrence over developmental periods) versus 2+ developmental periods (recurrence). |
| **Extent of exposure to different or repeating acts, by type of maltreatment** | Standardized average of endorsed items ranging from 0-10. | Standardized average of endorsed items ranging from 0-10. |

*Note.* Data were compiled from the final master file of the Québec Longitudinal Study of Child Development (1998-2018), Québec Government, Institut de la Statistique du Québec.
Within each developmental period: No= none or exposure at a single age point; Yes= exposure at more than one age point. Within lifetime: No= none or exposure at one developmental period; Yes=more than one developmental period.
<sup>a</sup>By childhood includes preschool and school-age.
<sup>b</sup>By adolescence includes preschool, school-age and adolescence.

### Descriptive statistics of child maltreatment indicators

Descriptive statistics outlined the frequencies and means of the child maltreatment indicators (i.e., presence by type, recurrence, chronicity, extent of exposure) at each developmental period and by the end of childhood and adolescence. As response rates varied across developmental periods, we compared participants with valid data to those present at inception on key early-life individual and family characteristics (e.g., externalizing symptoms, socio-economic status) according to their status of missingness. We then examined the concordance between the prospectively derived and retrospectively reported indicators of child maltreatment using Cohen’s Kappa. To quantify the extent of discordance between these indicators, a percentage bias (Atherton et al., 2008) was also calculated which refers to the proportional difference between those that were included versus the initial cohort (sample(by developmental period)% - total initial cohort%)/total initial cohort%).

## Results

### Number of included items

A total of 251 items, out of a total of 29,600 items from birth to 17 years, were included to derive indicators of child maltreatment. These items as well as their respective cut-offs and informants are presented in **Supplemental Table 2**. Most items enquire about exposure of intrafamilial maltreatment for which the indicated time window was within the past 6 or 12 months (e.g., “In the past 12 months…”), or since the beginning of the school year. From 5 months to age 17 years, 60.0% of items were reported by the mother, 12.7% of items were drawn from the interviewer’s observational reports of the home environment (between birth to 56 months), 12.3% by the child’s schoolteacher (starting when children reached formal schooling, i.e., 6 years old to 13 years) and 15.0% of items were child reported (starting at age 10 to 17 years). Notably, the number of items varied according to the maltreatment types and developmental periods. For example, psychological abuse was derived according to a varying number of items in preschool (*n*=16), school-age (*n*=2) and adolescence (*n*=2), whereas sexual abuse is measured solely in adolescence. Educational/supervisory neglect contains the most items (*n*=26 unique items) from birth to 17 years.

### Prospective prevalence rates of maltreatment indicators

#### Presence by maltreatment type

Prevalence rates for the types of child maltreatment are presented in **Table 3** within developmental periods and by the end of childhood and adolescence. Across all developmental periods, physical abuse varies from 16.3-21.8% while psychological abuse varies from 3.3-21.9%, emotional neglect from 20.4-21.6%, physical neglect varies from 15.0-22.3%, supervisory neglect from 25.8-44.9%, family violence from 4.1-11.2% and sexual abuse was present in 9.5% of the population in adolescence. Estimates by the end of adolescence (birth to 17 years) across all maltreatment types range from 16.5-67.3%.

**Table 3.** Prevalence estimates of probable childhood maltreatment indicators across developmental periods, the lifetime and retrospective reports (%, n)

|  | **Preschool**<br>(birth to 5<br>years) | **Childhood**<br>(6 to 12<br>years) | **Adolescence**<br>(13 to 17<br>years) | **By the end of**<br>**childhood** (birth<br>to 12 years)<sup>a</sup> | **By the end of**<br>**adolescence** (birth<br>to 17 years)<sup>b</sup> | **Retrospectively assessed**<br>**at age 23 years** (birth to<br>18 years) |
| --- | --- | --- | --- | --- | --- | --- |
| **Maltreatment types** |  |  |  |  |  |  |
| Physical abuse | 21.8(427) | 17.4(236) | 16.3(227) | 29.7(400) | 37.3(446) | 4.9(64) |
| Sexual abuse | - | - | 9.5(114) | - | - | 11.7(154) |
| Psychological abuse | 21.9(426) | 3.3(39) | 6.8(94) | 22.6(263) | 25.7(277) | 13.6(179) |
| Emotional neglect | 20.7(413) | 21.6(285) | 20.4(237) | 34.9(458) | 42.1(431) | 14.6(192) |
| Physical neglect | 15.0(299) | 22.3(199) | - | 30.3(270) | - | 2.5(33) |
| Supervisory/educational neglect | 25.8(508) | 44.9(562) | 36.5(497) | 55.2(683) | 67.3(715) | - |
| Family Violence | 11.2(209) | 8.5(99) | 4.1(44) | 16.0(184) | 16.5(150) | 4.2(56) |
| **Cumulative maltreatment<sup>c</sup>** | *n=1969* | *n=1221* | *n=1309* | *n=1207* | *n=964* |  |
| 0 | 36.6(720) | 40.7(497) | 46.3(606) | 22.0(266) | 15.9(153) |  |
| 1 | 32.7(643) | 34.5(421) | 32.6(427) | 30.6(369) | 31.6(305) |  |
| 2 | 18.5(364) | 16.2(198) | 13.1(172) | 21.9(264) | 25.6(247) |  |
| 3+ | 12.3(242) | 8.6(105) | 7.9(104) | 25.5(308) | 26.9(259) |  |
| **Cumulative maltreatment**<br>**(without supervisory/educational**<br>**neglect)<sup>d</sup>** | *n=1952* | *n=1129* | *n=1169* | *n=1111* | *n=1019* | *n=1323* |
| 0 | 44.3(865) | 58.4(659) | 64.2(750) | 35.0(389) | 33.2(338) | 69.7(922) |
| 1 | 33.5(653) | 29.9(338) | 23.5(275) | 31.4(349) | 34.9(356) | 18.6(246) |
| 2 | 15.1(139) | 8.9(101) | 8.8(103) | 20.6(229) | 20.5(209) | 5.8(77) |
| 3+ | 7.1(139) | 2.7(31) | 3.5(41) | 13.0(144) | 11.4(116) | 5.9(78) |

*Note.* Data were compiled from the final master file of the Québec Longitudinal Study of Child Development (1998-2018), Québec Government, Institut de la Statistique du Québec.
The number of items vary by maltreatment type and across each developmental period. See supplemental Table 2 for more information.
<sup>a</sup>By the end of childhood includes preschool and school-age.
<sup>b</sup>By the end of adolescence includes preschool, school-age and adolescence.
<sup>c</sup>Cumulative maltreatment by the end of adolescence (birth to 17 years) excludes sexual abuse and physical neglect as they are not available at all three developmental periods.
<sup>d</sup>Given the relatively high prevalence of supervisory/educational neglect, we also present cumulative maltreatment while excluding this category.

#### Cumulative maltreatment

Cumulative maltreatment across development periods and retrospectively is presented in **Table 3**. Given the high prevalence of supervisory/educational neglect (67.3% by adolescence) and unavailability of corresponding retrospective indicators, we also estimated cumulative maltreatment excluding supervisory/educational neglect. The occurrence of 0, 1, 2, and 3+ maltreatment types, excluding supervisory/educational neglect, was distributed as follows by the end childhood 35.0%, 31.4%, 20.6% and 13.0% and by the end of adolescence, 33.2%, 34.9%, 20.5%, and 11.4%, respectively.

#### Extended indicators of maltreatment

**Table 4** presents the recurrence of each type of maltreatment within and across developmental periods (i.e., chronicity). This indicator captures exposure to each type of maltreatment at more than one age point within a developmental period. Estimates of recurrence by the end of adolescence varied between 3.2-29.5% across the five types of maltreatment indexed at all three developmental periods (physical abuse, psychological abuse, emotional neglect, supervisory/educational neglect, family violence). As expected, our indicator of extent of exposure to different or repeating acts **(Table 5)**, both within and across developmental periods, was highly skewed, indicating that most children are not exposed to numerous maltreatment acts.

**Table 4.** Recurrence of probable child maltreatment (%, n)

|  | **Preschool**<br>(birth to 5 years) |  | **School-age**<br>(6 to 12 years) |  | **Adolescence**<br>(13 to 17 years) |  | **By the end of**<br>**childhood**<br>(birth to 12 years)<sup>a</sup> |  | **By the end of**<br>**adolescence**<br>(birth to 17 years)<sup>b</sup> |  |
| --- | --- | --- | --- | --- | --- | --- | --- | --- | --- | --- |
|  | No | Yes | No | Yes | No | Yes | No | Yes | No | Yes |
| **Physical abuse** | 92.0(1789) | 8.0(156) | 95.7(1266) | 4.3(57) | 98.0(1349) | 2.0(27) | 90.9(1223) | 9.1(122) | 88.0(1053) | 12.0(144) |
| **Sexual abuse** | - | - | - | - | 99.3(1175) | .7(8) | - | - | - | - |
| **Psychological abuse** | 96.0(1856) | 4.0(77) | 99.7(1180) | .3(4) | 99.2(1362) | .8(11) | 98.8(1148) | 1.2(14) | 96.8(1042) | 3.2(34) |
| **Emotional neglect** | 95.7(1892) | 4.3(85) | 96.6(1215) | 3.4(43) | 97.0(1083) | 3.0(34) | 92.2(1210) | 7.8(102) | 85.9(879) | 14.1(144) |
| **Physical neglect** | 97.1(1917) | 2.9(57) | 97.4(772) | 2.6(21) | - | - | 94.2(838) | 5.8(52) | - | - |
| **Supervisory/educational**<br>**neglect** | 94.1(1836) | 5.9(115) | 90.3(997) | 9.7(107) | 92.8(1219) | 7.2(95) | 83.8(1037) | 16.2(200) | 70.5(749) | 29.5(313) |
| **Family violence** | 97.9(1822) | 2.1(40) | 99.2(1145) | .8(9) | - | - | 97.5(1118) | 2.5(29) | 96.8(882) | 3.2(29) |

*Note.* Data were compiled from the final master file of the Québec Longitudinal Study of Child Development (1998-2018), Québec Government, Institut de la Statistique du Québec.
Within each developmetal period: No= none or exposure at a single age point; Yes= exposure at more than one age point. Within lifetime: No= none or exposure at one developmental period; Yes=more than one developmental period.
<sup>a</sup>By childhood includes preschool and school-age;
<sup>b</sup>By adolescence includes preschool, school-age and adolescence.

**Table 5.**
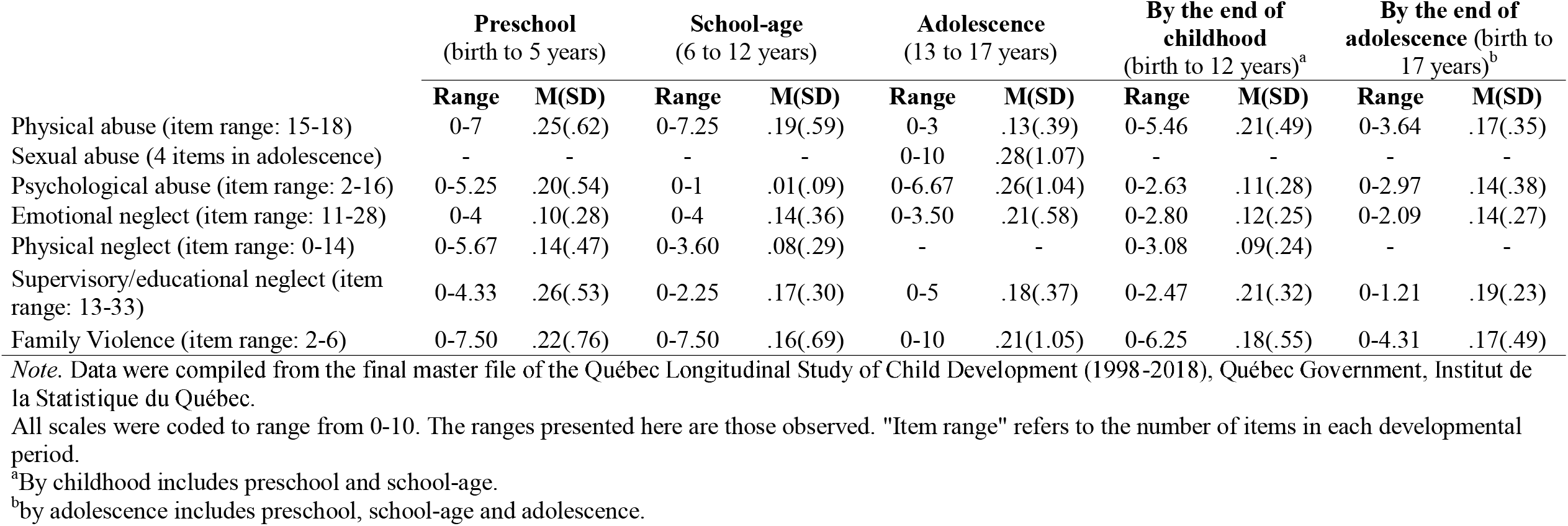
Extent of exposure to different or repeating acts of maltreatment by type across developmental periods.

|  | **Preschool**<br>(birth to 5 years) |  | **School-age**<br>(6 to 12 years) |  | **Adolescence**<br>(13 to 17 years) |  | **By the end of**<br>**childhood**<br>(birth to 12 years)<sup>a</sup> |  | **By the end of**<br>**adolescence** (birth to<br>17 years)<sup>b</sup> |  |
| --- | --- | --- | --- | --- | --- | --- | --- | --- | --- | --- |
|  | **Range** | **M(SD)** | **Range** | **M(SD)** | **Range** | **M(SD)** | **Range** | **M(SD)** | **Range** | **M(SD)** |
| Physical abuse (item range: 15-18) | 0-7 | .25(.62) | 0-7.25 | .19(.59) | 0-3 | .13(.39) | 0-5.46 | .21(.49) | 0-3.64 | .17(.35) |
| Sexual abuse (4 items in adolescence) | - | - | - | - | 0-10 | .28(1.07) | - | - | - | - |
| Psychological abuse (item range: 2-16) | 0-5.25 | .20(.54) | 0-1 | .01(.09) | 0-6.67 | .26(1.04) | 0-2.63 | .11(.28) | 0-2.97 | .14(.38) |
| Emotional neglect (item range: 11-28) | 0-4 | .10(.28) | 0-4 | .14(.36) | 0-3.50 | .21(.58) | 0-2.80 | .12(.25) | 0-2.09 | .14(.27) |
| Physical neglect (item range: 0-14) | 0-5.67 | .14(.47) | 0-3.60 | .08(.29) | - | - | 0-3.08 | .09(.24) | - | - |
| Supervisory/educational neglect (item range: 13-33) | 0-4.33 | .26(.53) | 0-2.25 | .17(.30) | 0-5 | .18(.37) | 0-2.47 | .21(.32) | 0-1.21 | .19(.23) |
| Family Violence (item range: 2-6) | 0-7.50 | .22(.76) | 0-7.50 | .16(.69) | 0-10 | .21(1.05) | 0-6.25 | .18(.55) | 0-4.31 | .17(.49) |

*Note.* Data were compiled from the final master file of the Québec Longitudinal Study of Child Development (1998-2018), Québec Government, Institut de la Statistique du Québec.
All scales were coded to range from 0-10. The ranges presented here are those observed. "Item range" refers to the number of items in each developmental period.
<sup>a</sup>By childhood includes preschool and school-age.
<sup>b</sup>by adolescence includes preschool, school-age and adolescence.

#### Concordance between retrospective and prospective maltreatment indicators

In comparison to child maltreatment prevalence based on prospectively collected data, retrospective measures of child maltreatment were much lower, ranging from 2.5-14.6% across all types of maltreatment (**Table 3**). **Table 6** shows that the concordance estimates between prospective (by the end of adolescence) and retrospective reports by types of maltreatment were small (.038 - .110), yet statistically significant (*p*_s_ = <.01), except for emotional neglect (*p*= .14). Of note, 29.9% (n=190) of individuals with any type of maltreatment documented from birth to 17 years using our prospective index subsequently reported maltreatment at age 23 years (kappa: .067, *p*= .003). The degree of concordance between prospective and retrospective cumulative maltreatment (0, 1, 2, 3+) was small but significant (kappa=.058, *p* = .001).

**Table 6.** Agreement between our prospective presence indicator (by the end of adolescence) and retrospective maltreatment.

|  | K | *p* |
| --- | --- | --- |
| Physical abuse | .075 | <.001 |
| Sexual abuse | .110 | <.001 |
| Psychological abuse | .110 | <.001 |
| Emotional neglect | .037 | .148 |
| Physical neglect | .057 | .002 |
| Supervisory/educational neglect | - | - |
| Family Violence | .060 | .009 |
| Any types | .067 | .003 |
| Cumulative maltreatment | .058 | .001 |

*Note.* Data were compiled from the final master file of the Québec Longitudinal Study of Child Development (1998-2018), Québec Government, Institut de la Statistique du Québec.
K = kappa estimate.
Prospective physical neglect by the end of childhood was used to estimate agreement (by the end of adolescence not available). Prospective sexual abuse "adolescence" was used to estimate agreement.
Supervisory/educational neglect is not included in prospective "any types", as it is not measured retrospectively.

#### Quantifying attrition and non-response

Due to attrition and non-responses, the sample sizes varied according to each maltreatment indicator. Participants with valid data for each derived indicator were compared to the initial cohort on key characteristics that have the potential to identity the most vulnerable participants, thus most likely to be lost to follow up. This comparison is expressed as percentage bias (Atherton et al., 2008), **Table 3 in Supplemental material**). Biases ranged from 0% (for internalizing and externalizing behaviors) to 36.36% (for maternal age at birth). Across all developmental periods and retrospective indicators, participants with missing data tended to be male (e.g., in school-age and retrospective reports), to be of non-Canadian descent (e.g., in adolescence), to be born to a mother younger than 20 years old (e.g., by the end of adolescence) or who reported higher levels of depressive symptoms (e.g., in adolescence), to have grown-up in a single-headed or blended family (e.g., by the end of adolescence) or in a family with a lower socioeconomic status (e.g., by the end of adolescence).

## Discussion

This article outlines our strategy to derive prospective indicators of maltreatment anchored in a developmental perspective using various time-relevant indicators of maltreatment (e.g., recurrence, chronicity), rarely assessed in the literature, especially in population-based cohorts. Using a systematic screening approach, child maltreatment experts retained a total of 251 items from an original pool of 29,600 available items. These items were used to derive five indicators: maltreatment presence and cumulative scores, as well as recurrence, chronicity, and the extent of exposure. By the end of adolescence (birth to 17 years), a little more than one in three children (37.3%) were exposed to probable physical abuse, 9.5% to probable sexual abuse (measured in adolescence only), 25.7% to probable psychological abuse, 42.1% to probable emotional neglect, 30.3% to probable physical neglect (preschool and school-age), 67.3% to probable supervisory/educational neglect and 16.5% to probable family violence. The concordance between prospective and retrospective maltreatment types were low in magnitude, but significant (except for emotional neglect).

### Comparing our prospective estimates with other prospective cohort estimates

Comparison of our prospective maltreatment indicators with other cohort estimates is challenging. To our knowledge, there are no other cohorts that have derived probable maltreatment using several indicators (i.e., type, cumulative, recurrence, chronicity, extent of exposure to different or repeating acts) according to a longitudinal and non-specific item approach (not specifically designed to assess maltreatment). The ALSPAC cohort adversity index (Houtepen et al., 2018) was derived using a similar general-item and cut-off dichotomization approach. A total of 136 prospective items were used to identify maltreatment defined by abuse or neglect and 43 items were used to identify maltreatment retrospectively. The prevalence rates in ALSPAC were somewhat comparable to ours: physical abuse (ALSPAC: 17.4% vs. QLSCD: 37.4%), sexual abuse (3.7% vs. 9.5%), emotional abuse (22.5% vs. 25.7%), emotional neglect (22.1% vs. 42.1%), and family violence (24.1% vs. 16.5%), with a trend for higher probable prevalence in our cohort (family violence being a notable exception). Specifically, our prevalence rates for physical abuse and emotional neglect are comparable to ALSPAC when considering the individual developmental periods, however, our rates derived by the end of childhood and adolescence are higher. In comparison to ALPSAC, the convergence of several varying items (e.g., in adolescence, our index contains information on physical abuse from a romantic partner) and informants (e.g., home observations) across developmental periods may lead to the increased detection of probable maltreatment. Additionally, our index spans more items (251 vs. 136 prospective maltreatment items) than ALSPAC and data is collected over fourteen timepoints across three developmental periods. Conversely, prospective physical abuse in ALSPAC was evaluated less frequently in adolescence. This may lead to missing prospective reports of intervening maltreatment. As such, it is important to consider that prevalence rates for maltreatment might be sensitive to the number and types of items, informants, and timing at which the information was sought. Notably, ALSPAC used prospective and retrospective maltreatment information interchangeably (i.e., physical abuse was deemed present whether reported prospectively or retrospectively). However, as prospective and retrospective maltreatment reports may identify different groups of individuals (Baldwin et al., 2019), it is now recommended to treat prospective and retrospective separately. Direct comparisons between our prospective prevalence rates and other cohorts (e.g., Environmental Risk Longitudinal Twin Study and the Dunedin Longitudinal Study) is difficult given that the approaches differ, however, our prevalence rates tend to be higher compared to cohorts that use specific item approaches (i.e., items that target maltreatment) (Newbury et al., 2018; Reuben et al., 2016).

### Comparing prospective and retrospective reports (concordance)

Concordance estimates between prospective and retrospective reports of maltreatment by type (.038-.110) demonstrate that those who report maltreatment experiences retrospectively are not necessarily the same individuals who are identified in prospective reports, which falls in line with the slight to fair agreement found in previous studies (Baldwin et al., 2019). Relatedly, previous studies have found stronger associations between retrospective reports of child maltreatment and mental health later in life (Danese & Widom, 2020), which may point to potential bias in self-reports affected by current mental states and due to the same-informant and same methods shared variance between these measures. Notably, however, the studies included in Baldwin et al. (2019)’s analysis contained a variety of prospective report types (e.g., self, parent, medical records), but mainly reports from Child Protective Services. Conversely, our prospective estimates are based on multiple informants through questionnaire format (and home observations). In the QLSCD, the retrospective report was solely based on self-report questionnaire items, whereas those in Baldwin et al. (2019) included interviews in addition to self-report questionnaires. According to Baldwin et al. (2019), the concordance between prospective and retrospective reports was higher in studies that used interview versus questionnaires in retrospective self-reports, which may indicate that our estimate of concordance is conservative.

Nonetheless, concordance estimates have been found to be low, thus, prospective and retrospective reports of maltreatment should be kept separate. However, future cohorts may consider collecting both prospective and retrospective maltreatment data to further explore differential associations.

### Methodological considerations

Our study had the following strengths. Information was collected from four types of informants (parents, teachers, the target child, and interviewer’s observations), allowing us to capture multiple perspectives and schemes of reference. Further, given the longitudinal nature of the QLSCD cohort, comprising data collected at 14 time points, our indicators offer insight into the probable presence of maltreatment occurring at different developmental periods in early life (preschool, school-age and adolescence). As such, our study provides opportunities to examine more often the role of time-varying characteristics of maltreatment (other than presence of maltreatment), including chronicity and recurrence, by providing researchers a blueprint guiding their creation in longitudinal cohorts that did not explicitly measure various types of maltreatment. The definitions selected to guide the maltreatment experts for item selection reflected the Québec Youth Protection Act and supporting resources (*Grounds for Reporting a Situation*, 2022; Québec, 2021). These definitions generally align with conventional definitions and categorizations of maltreatment, such as the Centers for Disease Control and Prevention report (Arias et al., 2008), and the United Kingdom government report on Working Together to Safeguard Children (Government, 2018). Notably, we used a rigorous screening process to extract relevant items in collaboration with experts in child development and maltreatment. The standardized sum of endorsed items was highly skewed, representing more conservative thresholds to determine the *probable* presence of child maltreatment. Further, bias was minimized as the maltreatment experts decided on the cut-offs for each of the items prior to analyzing prevalence rates of the derived variables and engaged in discussions to minimize subjective risk.

However, our study also has limitations that need to be considered when interpreting the results. First, the pool of items available in the QLSCD was not originally designed to assess maltreatment. While we included a wide range of potential harmful behaviors to derive our indicators (e.g., presence), such as “I have shaken my baby/twin when he/she was particularly fussy” and “there was more than one incident involving physical punishment during the visit” (for physical abuse), no individual item alone indicates a definitive presence of maltreatment. Second, we were unable to derive an indicator of severity as based on the relative frequency of occurrence of each item. For instance, while physical abuse is measured in terms of “hitting” and “shaking”, other severe forms are not available, such as “kicking or “chocking”. Moreover, severe cut-off scores were selected for each item as indicative of probable maltreatment, depending on the developmental period (e.g., the cut off for “in the past 6 months, your parents hit you or threaten to do so” was “often” when this item was measured in adolescence). Instead, we opted to derive the indicator “extent of exposure to different or repeating acts” as reflective of the relative extent of exposure to each type of maltreatment. However, this indicator captures indistinctively a) repeated acts (e.g., same items present at two different time points) and b) the variety of acts within a given type (e.g., two different items within the same time point).

Third, similarly to all measurement methods, there is a risk of over- and under estimation of maltreatment types as based on social desirability and parents’ mental states, for instance, and we cannot ascertain whether the prevalence rates are “true” representations of maltreatment in the QLSCD (Denholm et al., 2013; Fallon et al., 2010; Mathews et al., 2020). Fourth, the generally high prevalence of supervisory/educational neglect may reflect a higher number of items in comparison to other types, despite using a stringent cut off for each item (e.g., the response “often” for “in the past 12 months, how often did he/she see television shows or movies that have a lot of violence in them?” was coded as “probable maltreatment”). This finding is nevertheless consistent with a cross-sectional Québec population-based study that evaluated supervisory neglect using the short version of the Parent-Report Multidimensional Neglectful Behavior Scale, which found this type of maltreatment to have the highest annual prevalence rates (e.g., 24% for children 5-9 years) (Clément et al., 2016). On a related point, psychological abuse and sexual abuse may have been underestimated given the detection of less relevant items. The screening for sexual abuse was limited to late adolescence and covered sexual abuse with a romantic partner only (i.e., experiences that may have occurred in infancy or childhood, as well as in other contexts may have been missed). It is also important to consider that the family violence subtype combines items that reflect instances of family violence (without the guarantee that the child witnessed the violence), and most items only evaluate past 12-month trauma exposure at 41 and 45-56months and 5, 6, 8, 10, 12 and 13 years, which may have missed intervening trauma. Fifth, although our prevalence rates are generally consistent across developmental periods (preschool, school-age and adolescence; 21.8%, 17.4% and 16.3% for physical abuse, respectively), comparison across developmental periods is not without bias, as discussed previously. Specifically, there is the possibility that the prevalence rates vary depending on the number of items used to derive the variables. For instance, to derive psychological abuse in preschool, there are 15 items, whereas there were only 2 items to derive school-age exposure. As such, comparison across developmental periods should be examined cautiously. Sixth, there are limitations regarding the representativeness of the cohort.

Indigenous youth were excluded, yet they are more likely to report maltreatment compared to non-Indigenous youth (Government of Canada, 2017). Differential longitudinal attrition occurred among the most vulnerable participants (i.e., biases existed in our samples compared to the initial cohort) and as expected, there is a general increase in bias as the cohort ages. For instance, individuals from a lower socioeconomic status and blended families were increasingly underrepresented, however, the extent of biases are minimal. Finally, the retrospective measure of child maltreatment available in the QLSCD is based on a checklist of only six items and does not provide detailed information on supervisory neglect, as well as important characteristics of maltreatment such as timing and chronicity.

### Future directions

The method used to derive our indicators of child maltreatment offers a relatively novel approach for capturing probable maltreatment in population-based cohorts. Future cohorts may consider undertaking a similar approach to broaden research investigations through the consideration of extended maltreatment characteristics that are often difficult to capture. As a next step, we will examine the validity of this approach, and the indicators that resulted from it, by investigating and comparing the prospective and retrospective associations with mental health outcomes, such as depression, and suicidality and early-life correlates such as family socioeconomic status and dysfunction. Child maltreatment is global problem with consequences at the societal and individual level. Our index offers a pragmatic and prospective approach to detecting child maltreatment for research purpose in datasets where it is not directly assessed.

### Maltreatment types Definitions

Physical abuse A situation in which the child is the victim of bodily injury or is subjected to unreasonable methods of upbringing by his parents or another person, and the child’s parents fail to take the necessary steps to put an end to the situation.

## Data Availability

All data produced in the present work are contained in the manuscript

https://www.iamillbe.stat.gouv.qc.ca/default_an.htm

## Acknowledgement

The QLSCD is conducted by the Institut de la Statistique du Québec. Data were compiled from the final master file of the Québec Longitudinal Study of Child Development (1998–2018), Québec Government, Institut de la Statistique du Québec.

## Notes

**Disclosure:** Ms Scardera reported receiving a doctoral award from the Social Sciences and Humanities Research Council (SSHRC). Dr Langevin is supported by a Chercheur-Boursier Award from the Fonds de recherche du Québec - Santé. Dr. Collin-Vezina holds the Nicolas Steinmetz and Gilles Julien Chair in Community Social Pediatrics. Maude Comtois-Cabana received doctoral fellowships from the SSHRC and the Fonds de recherche du Québec – Société et culture (FRQSC). Dr Pinto Pereira was supported by a UK Medical Research Council Career Development Award (ref: MR/P020372/1). Dr Ouellet-Morin holds a Canada Research Chair in the Developmental Origins of Vulnerability and Resilience. Dr Geoffroy holds a Canada Research Chair in Youth Mental Health and Suicide Prevention. No other disclosures were reported.

**Funding/Support:** The Québec Longitudinal Study of Child Development was supported by funding from the Ministère de la Santé et des Services Sociaux, Ministère de la Famille, and Ministère de l’Éducation and Ministère de l’Enseignement Supérieur (Québec Ministries), the Lucie and André Chagnon Foundation, the Institut de Recherche Robert-Sauvé en Santé et en Sécurité du Travail, the Research Centre of the Sainte-Justine University Hospital, the Ministère du Travail, de l’Emploi et de la Solidarité Sociale, and the Institut de la Statistique du Québec. Additional funding was received from the Canadian Institutes of Health Research awarded to Dr. Geoffroy.

### Competing Interest Statement

The authors have declared no competing interest.

### Funding Statement

Funding/Support: The Québec Longitudinal Study of Child Development was supported by funding from the Ministère de la Santé et des Services Sociaux, Ministère de la Famille, and Ministère de l'éducation and Ministère de l'Enseignement Supérieur (Québec Ministries), the Lucie and André Chagnon Foundation, the Institut de Recherche Robert-Sauvé en Santé et en Sécurité du Travail, the Research Centre of the Sainte-Justine University Hospital, the Ministère du Travail, de l'Emploi et de la Solidarité Sociale, and the Institut de la Statistique du Québec. Additional funding was received from the Canadian Institutes of Health Research awarded to Dr. Geoffroy.
Role of the Funder/Sponsor: The funder had no role in the design and conduct of the study; collection, management, analysis, and interpretation of the data; preparation, review, or approval of the manuscript; and decision to submit the manuscript for publication.

### Author Declarations

The QLSCD data collection is conducted by the Institut de la Statistique du Québec. All the data collected and presented in this study has been approved by ethical committees of Institut de la Statistique du Québec and the CHU Sainte-Justine Hospital Research Centre. The 2021 Special Round data collection (23 years) was also approved by the Douglas Research Center Ethics Committee and by the CHU Ste-Justine research ethics committee.

